# A randomised control trial investigating the effect of improving the cleaning of shared medical equipment on healthcare-associated infections (The CLEEN study): Statistical Analysis Plan

**DOI:** 10.1101/2023.12.20.23300169

**Authors:** Nicole White, Allen Cheng, Katrina Browne, Philip Russo, Andrew Stewardson, Maham Amin, Kirsty Graham, Jennie King, Peta Tehan, David Brain, Maria Northcote, Brett Mitchell

## Abstract

This document outlines the Statistical Analysis Plan for the CLEaning and Enhanced disiNfection (CLEEN) study. The CLEaning and Enhanced disiNfection (CLEEN) study is a stepped wedge cluster randomised trial evaluating the role of enhanced cleaning and disinfection of shared medical equipment as part of hospital infection prevention and control programs. The study is preregistered with the Australian and New Zealand Clinical Trials Registry (ACTRN12622001143718) and is funded by the National Health and Medical Research Council (GNT2008392). The full study protocol used to inform the Statistical Analysis Plan has been published.^1^ A signed copy of the Statistical Analysis Plan is available on request from the corresponding author (BM).

## 1.0 Administrative information

### 1.1 Study identifiers

- Protocol: Research protocol version 1.2, 20/01/2023.
- Australia New Zealand Clinical Trial Registry (ACTRN12622001143718).
- NHMRC grant funding: NHMRC Emerging Leadership Investigator grant (Prof Brett Mitchell), (GNT2008392).

### 1.2 Revision history

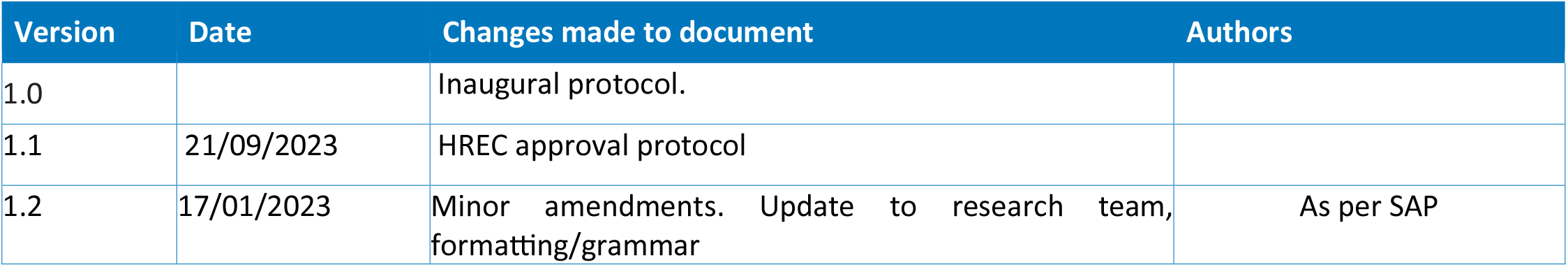

### 1.3 Contributors to the statistical analysis plan

#### 1.3.1 Roles and responsibilities

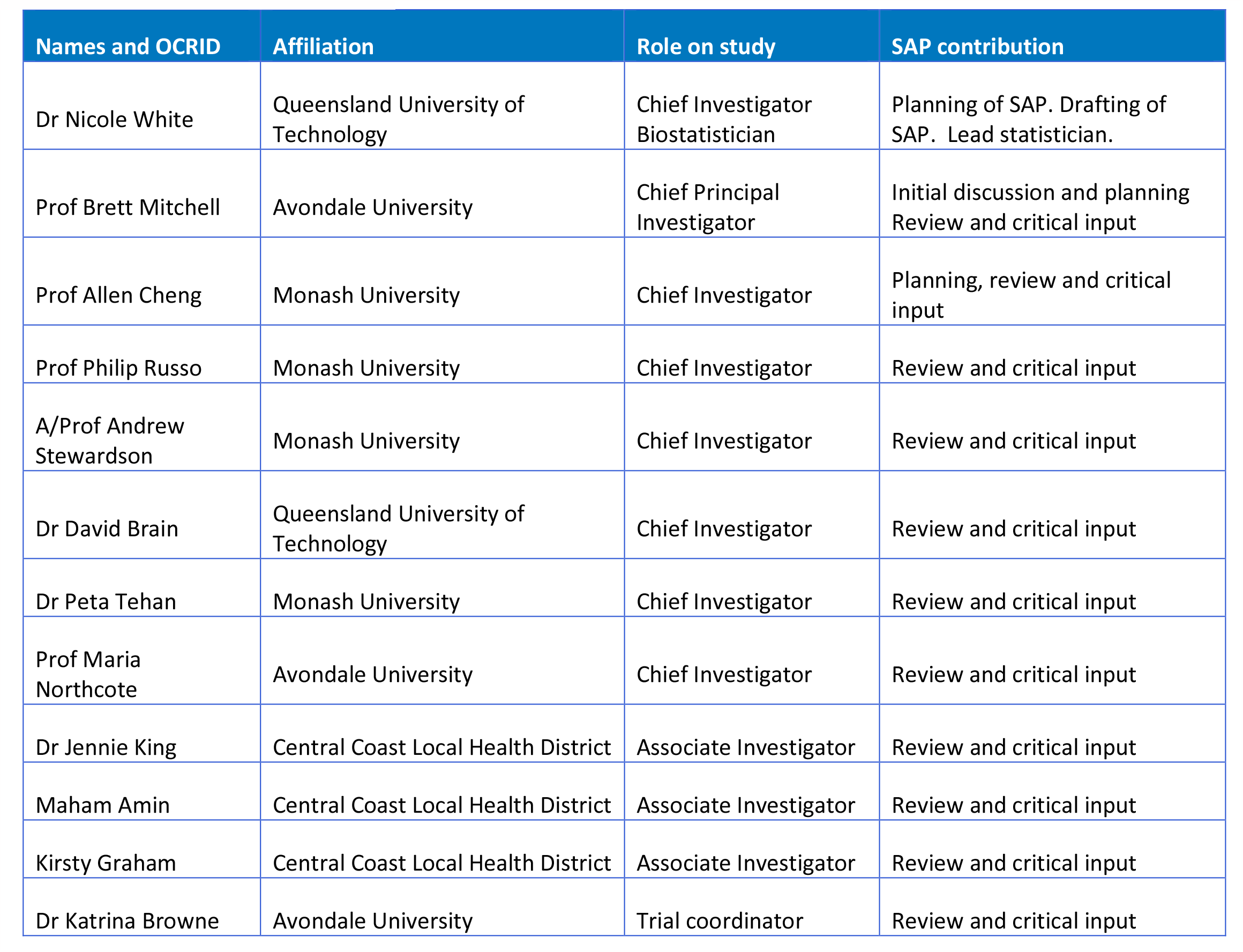

## 2.0 Study synopsis

The CLEaning and Enhanced disiNfection (CLEEN) study is a stepped wedge cluster randomised trial evaluating the role of enhanced cleaning and disinfection of shared medical equipment as part of hospital infection prevention and control programs.

The intervention to be evaluated is an evidence-based approach combining staff training, auditing, and feedback to environmental services staff on cleaning and disinfection practices.

### Study design

The trial will be conducted over 36 weeks in 10 wards within a large Australian tertiary hospital. The stepped wedge design will consist of five clusters, where each cluster comprises two wards. All clusters will start in the control condition, and one cluster will transition to the intervention condition every six weeks. Once a cluster is exposed to the intervention condition, it will remain exposed until the end of the trial. No transition period is assumed. Treatment sequences by cluster and ward are shown in Figure 1.

**Figure 1.**
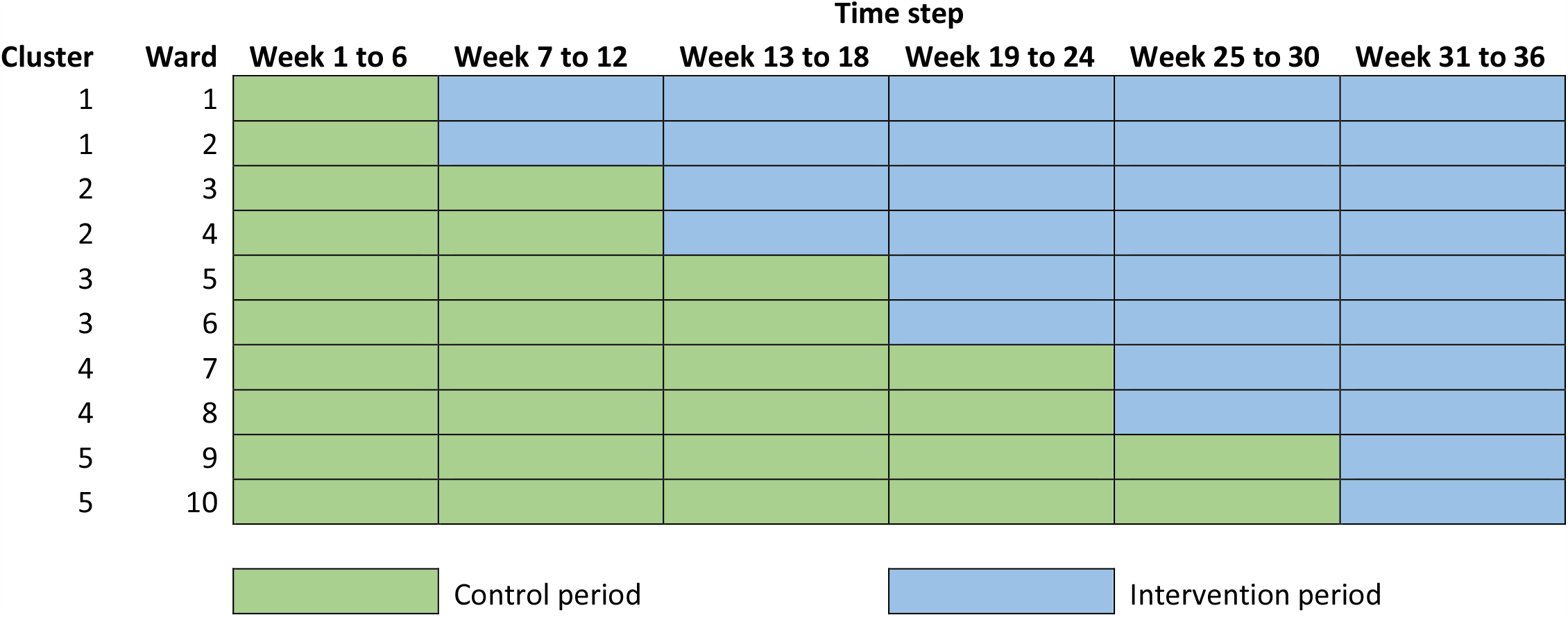
Study design.

### 2.1 Study objectives

#### 2.1.1 Primary objective

To evaluate the clinical effectiveness of enhanced cleaning and disinfection of shared medical equipment on healthcare-associated infection rates.

The hypothesis to be tested is that exposure to the CLEEN intervention will reduce the fortnightly proportions of healthcare-associated infections in participating wards, compared with pre-intervention incidence in the same participating wards.

#### 2.1.2 Secondary objectives

To evaluate changes in the thoroughness of shared medical equipment cleaning in participating wards before and after intervention exposure.

The hypothesis to be tested is that exposure to the CLEEN intervention is associated with an increase in the proportion of shared medical equipment that were successfully cleaned.

### 2.2 Patient population

#### 2.2.1 Inclusion criteria

- Data from all patients (>18 years old) who are admitted to one of the 10 wards on the day of the point prevalence survey
- All patients admitted to the ward before or at 08:00hrs on the first survey day and not discharged from the ward at the time of the survey will be eligible and will be evaluated for a HAI.

#### 2.2.2 Exclusion criteria

- Patients transferred in or out after 08:00hrs, will not be included. Patients will be excluded if they are under 18 years of age or are due to have same day treatment or surgery.

### 2.3 Outcomes

#### 2.3.1 Primary outcome

The primary outcome is confirmed cases of healthcare-associated infection. A healthcare-associated infection is defined as an infection that is acquired as a direct or indirect result of healthcare and is confirmed to have been acquired more than 48 hours after admission to the healthcare facility or associated with a recent admission or procedure. The determination of a healthcare-associated infection will be undertaken through an algorithm applying the healthcare-associated infection definitions in the European Centre for Disease Prevention and Control (ECDC) protocol, version 5.3._2_ Data on each HAI identified will be consistent with the ECDC protocol. Examples of infections included, but not limited to, include, blood stream, surgical site infection, pneumonia, urinary tract and soft tissue.

The primary outcome will be measured every two weeks using a standardised, validated point prevalence survey. The atribution of a healthcare-associated infection to a participating ward will be determined using a 48-hour time frame (2 days for practical reasons), i.e., the infection symptom onset must occur more than 48 hours after admission to the ward, for it to be atributable to the ward. If a patient is transferred to a ward and a HAI is identified within 48 hours of transfer, it will be atributed to the previous ward. Atribution based on the timing of a cluster’s transition from control to intervention will follow the rules listed below:

i. If a patient is admitted to a ward before the start of the control period of the trial and acquires a healthcare-associated infection less than 48 hours from the start of the control period, the infection will be excluded.
ii. If a patient is admitted to a ward before the start of the intervention period, and acquires a healthcare-associated infection in the same ward after the start of the intervention period:
  a. The infection will be included in the final week of the control period if found within 48 hours from the start of the intervention period.
  b. The infection will be included in the relevant week of the intervention period if found more than 48 hours from the start of the intervention period.
iii. If a patient is admitted to a ward during the intervention period, and acquires a healthcare-associated infection in the same ward after the trial end date:
  a. The infection will be included in Week 36 if found within 48 hours of the trial end date.
  b. The infection will be excluded if found more than 48 hours after the trial end date.

An interrater reliability assessment will be independently undertaken to validate the accuracy of HAI determinations.

#### 2.3.2 Secondary outcome

The secondary outcome is thoroughness of shared medical equipment. Shared medical equipment is defined as medical equipment used by patients admitted to the ward or by ward staff as part of healthcare delivery. Shared medical equipment included in this study are: Intravenous drip stands/poles, infusion pumps, mobile blood pressure machines, computer on wheels, commodes, blood glucose machines, medication trolleys, surgical trolleys, bladder scanners, patslides, resuscitation trolleys and mobility equipment including walkers, wheelchairs, pick up frames and rollator frames. The thoroughness of cleaning will be measured through fortnightly audits on study wards. Each audit will assess a random subset of shared medical equipment used in each study ward. Audits will commence in Week 2 of the trial.

### 2.4 Intervention

The CLEEN study will evaluate an intervention bundle consisting of four evidence-based components: education on cleaning technique, additional time dedicated to cleaning and disinfection of shared medical equipment by dedicated environmental services staff, auditing the thoroughness of shared medical equipment cleaning and feedback of audit results to staff. Full details of the intervention are available in the published study protocol (DOI: 10.1186/s13063-023-07144-z).^1^

All study wards will receive the intervention based on randomised treatment allocations in Figure 1.

### 2.5 Randomisation and blinding

All ten wards were randomised to treatment sequences after all wards had been recruited and before the start of the control period. Randomisation was completed in R using a set seed, where each ward was assigned a random number from 1 to 10 as per the treatment sequences in Figure 1. The trial statistician who is not located or affiliated with the trial hospital completed the randomisation.

The researcher(s) collecting primary and secondary outcome data was blinded to the intervention/treatment sequences. Results of the audit process for secondary outcome measurement were not made available to wards during the control period. Trial cleaning staff were blinded to intervention until they commence cleaning on the ward. The researcher(s) collecting primary and secondary data were blinded to the intervention/treatment sequences for the entirety of the study, until data analysis was complete.

The trial statistician, principal investigator and research coordinator will be unblinded to the treatment sequences. The final dataset will provide ward and cluster identifiers as anonymous codes for analysis and reporting.

### 2.6 Sample size

The study design was powered to detect a statistically significant, relative reduction of 35% in the primary outcome. The baseline HAI prevalence was assumed to be 11%, based on published data.^3,4^ Calculations for the study design in Figure 1 were performed in the Shiny CRT calculator [htps://clusterrcts.shinyapps.io/rshinyapp/], assumed a two-sided alternative hypothesis and a 5% level of statistical significance. Additional parameters to calculate the minimum sample size were an inter-cluster correlation of 0.3, and a coefficient of variation of 0.65. To achieve an expected power of 0.8 (80%), the minimum cluster size is 132 patients per time step, or 66 patients per ward per time step. The total minimum sample size based on these parameters is n = 3,960. Varying the intra-cluster correlation between 0.2 and 0.5 produced minimum sample sizes per cluster-time step of 94 (47 per ward) to 150 (75 per ward).

## 3.0 Statistical analysis

### 3.1 General principles

This plan outlines the analysis of HAI prevalence (primary outcome) and the thoroughness of shared medical equipment cleaning (secondary outcome). The planned cost-effectiveness analysis described in the published study protocol will be described in a separate analysis plan.

All statistical analyses and graphics will be completed in R version 4.0.5 or higher. R packages used for analysis will be reported in a software statement in the final analysis report. Analysis code will be made publicly available on GitHub (https://github.com/nicolemwhite).

Initial data cleaning will be completed by the research coordinator. The final dataset will be sent to the trial statistician in .csv or .xlsx format. Further data processing by the trial statistician will include the merging of data files by unique identifier(s) (where applicable) and checks of date-time variables against control and intervention period definitions as per the study protocol. Systematic data quality checks (e.g., negative numbers of cases, implausible dates) will be run to identify potential data entry errors for resolution before analysis commences.

Analyses will be reported in accordance with the Consolidated Standards of Reporting Trials (CONSORT) extension for stepped wedge cluster randomised trials (https://www.bmj.com/content/363/bmj.k1614)

There will be no interim analysis as we do not expect there to be any negative effects from this study.

### 3.2 Summary of changes

#### 3.2.1 Subgroup analyses

The primary outcome includes all confirmed cases of HAIs. The following subgroup analysies will be performed:

1. Analysis of bloodstream infection (BSI), urinary tract infection (UTI), pneumonia (PN) and surgical site infection (SSI) combined will occur. Subgroup analyses will independently re-run the primary outcome statistical model for each of these type of infection (BSI, UTI, PN, SSI). Model convergence will depend on the numbers of each HAI type reported across study weeks. The final analysis report will include a statement on which subgroup models did not converge (if any) AND
2. Analysis of all confirmed cases of HAI (same as primary outcome) with the exception of cases of COVID-19. AND
3. Analysis of confirmed cases of HAI (same as primary outcome) with the exception of cases of eye, ear, nose, mouth and throat (EENT) infections. Pilot work, undertaken prior to study commencement to test data collection processes, suggested this infection appeared to have a higher prevalence than in previously reported studies.

#### 3.2.2 Sensitivity analyses

Primary and secondary outcome models will estimate the average within-ward change associated with intervention exposure. A leave-one-ward-out analysis will be conducted to examine changes in estimated intervention effectiveness. Results reported for each model run will be the fixed effect for the intervention (Estimate with 95% confidence interval using model-based standard errors), and Cook’s distance as a measure of influence. The estimated study time trend (Estimate with 95% confidence interval using model-based standard errors) will also be reviewed to determine the sensitivity of findings to events unrelated to the intervention (e.g., ward outbreak).

The study protocol states that the primary and secondary outcome will be modelled by a Binomial Generalised Linear Mixed Model (GLMM) with logit link function. Results from this analysis will therefore be interpreted as odds ratios. Sensitivity analyses will refit models with log and identity link functions in place of the logit link function. Results provide estimates of intervention effectiveness as a risk ratio (log link) and risk difference (identity link). For different link functions, there is a possibility that the model will not converge due to sparse data. Analyses will consider the logit link first, as per the published protocol, followed by the identity link and the log link.

Model specification of study time will be considered as a categorical fixed effect, and a linear fixed effect. The appropriateness of categorical and linear effects will be assessed by residual diagnostics. In the event the both effects do not provide an adequate fit to the data, a first-order fractional polynomial for study time will be tested. It is possible that one or more models do not converge if data are sparse. Similarly, it is possible that the linear trend does not adequately reflect known events during the study that are unrelated to the intervention, for example, an outbreak across multiple wards or time periods.

In the final analysis report, results for converged models will be reported; a statement on which models did not converge will be included. Any outbreaks in study wards will also be reported to contextualise the choice of study time trend.

### 3.3 Blind review

A blinded review is planned (with the exception of the lead Biostatistician and Lead Chief Investigator) until there is agreement of the final presentation of results.

### 3.4 Data sets to be analysed

We will use an intention-to-treat analysis approach to analysis.

#### 3.4.1 Subject disposition

A CONSORT flowchart will be used to summarise the number of eligible admissions and exclusions based on outcome definitions in Section 2.3. Flowchart information will align with guidance provided by the cluster randomised trial extension to the CONSORT reporting checklist. ^5^

#### 3.4.2 Patient characteristics and baseline comparisons

Cohort characteristics will be summarised descriptively at the hospital- and ward-levels, stratified by study period (control, intervention exposure). Age-sex distributions will be presented graphically. Other descriptive statistics will be presented in tabular format.

Continuous variables will be summarised as means with standard deviations of medians with lower and upper quantiles. Categorical variables will be summarised as frequencies (numerator and denominator) with percentages. Reporting the denominator will reflect the presence of any missing data.

All patient characteristics will be reported as collected without transformation (e.g., no dichotomization of continuous variables). There will be no hypothesis testing of differences in baseline characteristics between study periods.

### 3.5 Compliance to study intervention(s)

We will report on the estimated compliance with the intervention descriptively (i.e. percentage of wards that received three hours of cleaning each day).

### 3.6 Analysis of the primary outcome

#### 3.6.1 Main analysis

The primary outcome is the total fortnightly HAIs identified divided by the number of at-risk patients per ward. The primary outcome will therefore be analysed as a proportion. The analysis will estimate the within-ward change in the primary outcome before and after intervention exposure using a generalized linear mixed model (GLMM). The model will specify a logit link function to model the association between the expected outcome and intervention exposure after adjusting for clustering. A random effect per ward will be specified to account for correlation arising from repeated measurements over time.

Fixed effects are intervention exposure and study time. Study time will be modelled in two ways to examine the nature of background trends in the primary outcome. First, study time will be modelled as a categorical variable, where each level is a study week. Second, a linear effect for study week will be considered. It is possible that specifying study time as a categorial variable is not possible if data are sparse. If the model with study time defined as a categorical variable specification does not converge, the linear effect will be reported. In the event that a linear effect provides a poor fit to the data based on residual diagnostics, a first-order fractional polynomial will be tested (see Section 3.2.2). Intervention exposure will be defined as a categorical variable (no = 0, yes = 1). Intervention exposure will be coded as no for the entire control period and week 1 of the intervention period. This 1-week delay will reflect study wards becoming familiarized with the enhanced cleaning procedures.

GLMM results will be reported for all fixed effects as odds ratios with 95% confidence intervals. The relative percentage change in outcomes will be reported alongside odds ratios. Model-based bootstrapped estimates for control and intervention estimates, after adjusting for study time and clustering, will be reported. The observed intra-cluster correlation and coefficient of variation will be reported to inform future studies.^6^ Sensitivity analyses will refit models with log and identity link functions in place of the logit link function (see Section 3.2.2).

Model assessment will consider the form of GLMM link function, over-dispersion and the comparison of model fit with generalized estimating equations. The appropriateness of the GLMM link function will be assessed by comparing the fit of the logit link function with alternative link functions. Model fits will be compared using Akaike’s Information Criterion (AIC), pending model convergence. Lower AIC values indicate improved model fit – if the AIC difference between two models is similar (<2), the model with fewer parameters will be preferred. The presence of over-dispersion will be assessed by residual diagnostics.

We will undertake sensitivity analyses to determine the possibility of a delayed intervention effect of longer than 1 week, the influence of each ward on model estimates and the effect of the intervention on the most common and serious HAIs—pneumonia, urinary tract infections, bloodstream infections and surgical site infection. The delayed intervention effect will be modelled at 2 and 4 weeks after each ward’s intervention start date. The influence of each ward will be examined using a leave-one-ward-out analysis examining changes to the intervention effect and Cook’s distances (see Section 3.2.2).

Based on data collection instruments, we do not anticipate any missing data for the primary outcome.

### 3.7 Analysis of secondary outcomes

Secondary outcome data from fortnightly cleaning audits will be analysed using a binomial generalized linear mixed model with a logit link function. The GLMM dependent variable is the number of pieces of shared medical equipment that were audited as ‘cleaned’ divided by the total number audited per audit occasion. A random intercept will be included for each ward. The effect of the intervention will be tested in three ways: a binary intervention effect, to model an immediate improvement in cleaning; a linear intervention effect, defined as weeks after each ward’s intervention start date, to model a more gradual improvement over time; and a combined binary–linear intervention effect. As per the analysis of the primary outcome, the intervention effect will switch from ‘no’ to ‘yes’ 1 week after the start of the intervention period.

The following sensitivity analyses as per the primary outcome analysis will be completed: leave-one-ward-out analysis, specification of the study time effect, GLMM link functions, delayed intervention effect.

We anticipate and hope to prevent missing data through our approach to data collection. We will report details of missing data, including differences between participant groups, paterns of missingness and the reasons for missing data. Our findings will include a discussion of the implication of missing data on interpreting the results. Where an individual participant’s data is missing and we cannot accurately determine whether they have HAI or not, their data will be excluded from analysis (including denominator data for the purpose of determining prevalence).

## Data Availability

All data produced in the present study are available upon reasonable request to the authors

## List of abbreviations

HAI: Healthcare-associated infection
GLMM: Generalised linear mixed model
AIC: Akaike’s Information Criterion
ECDC: European Centre for Disease Prevention and Control
CLEEN: CLEaning and Enhanced disiNfection
BSI: Bloodstream infection
UTI: Urinary tract infection
PN: Pneumonia
SSI: Surgical site infection
EENT: Ear, eye, nose, mouth and throat

